# Risks and challenges in COVID-19 infection prevention and control in a hospital setting: perspectives of healthcare workers in Thailand

**DOI:** 10.1101/2022.04.21.22274131

**Authors:** Monnaphat Jongdeepaisal, Puri Chunekamrai, Rapeephan R Maude, Richard J Maude

## Abstract

In hospital settings, awareness of, and responsiveness to, COVID-19 are crucial to reducing the risk of transmission among healthcare workers (HCWs) and protecting them from infection. Healthcare professionals can offer insights into the practicalities of infection prevention and control measures and on how the protective equipment and training could best be delivered during the pandemic. This study aimed to inform the development of future recommendations to optimise compliance with appropriate use of these measures, and to improve the guidance to reduce their risk of the disease. Drawing on in-depth interviews with HCWs in a hospital in Thailand, several factors influence the use of multiple prevention measures: concerns about infection, availability of the equipment supply, barriers to work performance, and physical limitations in the hospital setting. Setting a ventilated outdoor space for screening and testing, and interaction through mobile technology, were perceived to reduce the transmission risk for staff and patients. Adequate training, clear guidelines, streamlined communications, and management support are crucial to encourage appropriate use of, and adherence to, implementation of infection prevention and control (IPC) measures among HCW. Further study should explore the perceptions and experience of health professionals in local health facilities and community-based workers during the pandemic, particularly in resource-limited settings.

## Background

With the multiple waves of COVID-19 transmission, health care workers (HCWs) play a central role in saving lives. These professionals are the driving force to achieving effective clinical management of patients during the pandemic. They are also among the population with highest risk of infection with COVID-19 due to their contacts with high-risk individuals and working environment (1). Of the 3.45 million global deaths due to COVID-19 reported to WHO from January 2020 to May 2021, only 6,643 were among HCWs with the true figure estimated to be much higher at around 80,000-180,000 (2). In Thailand, suspected cases of infection with emerging infectious disease caused by SARS-CoV-2 were identified during early January 2020 (3). In April 2020, 102 HCWs, or 4 percent of total cases at the time, were infected with COVID-19; 28 of which had been providing direct care to patients, six had close contact with other HCWs, and one had been conducting screening and triage (4). Indigenous cases of COVID-19 were found to be minimal during the first wave (5); however, the larger second wave occurred from mid-December 2020 onwards (6). Subsequent waves of the pandemic in 2021, during which the number of cases increased in Bangkok and across the country (7), represented a significant rise in the burden for hospitals, with over 600 healthcare workers reported to be infected with COVID-19 despite receiving two doses of Sinovac vaccine (8). Most recently, in early 2022, there has been a rise in cases and staff absences due to the Omicron variant with potential for rapid spread including in healthcare settings. With the ongoing uncertainty and possibilities of new outbreaks in the future, COVID-19 remains a dangerous infectious disease, particularly for HCWs involved in managing patients with the disease.

In hospital settings, awareness of, and responsiveness to, COVID-19 are crucial to reducing the risk of transmission among HCWs and protecting them from infection. To protect HCWs, WHO’s guideline for COVID-19 IPC in healthcare settings recommends use of personal protective equipment (PPE), hand hygiene and IPC training (9). To inform the identification of risk factors for COVID-19 infection among HCWs, WHO has investigated the extent of infection in health care settings and risk factors for infection among HCWs (10). A survey of healthcare workers in 57 countries found that an overall high level of awareness and preparedness among HCWs who received training courses during the first wave of the pandemic, with variation regarding to gender, type of HCWs, and prior experience with outbreaks (11). A knowledge, awareness, and practice (KAP) assessment was conducted in Thailand to assess and improve knowledge, attitudes and practice among HCWs and the general public (12). To optimise compliance with appropriate use of PPE and IPC training, additional information is needed to understand the experience of HCWs during the pandemic and their opinions on how the PPE and IPC training could best be delivered.

Considering the ongoing need for protection from COVID-19 and the high level of risk among healthcare professionals, it is critical to explore the views and perceptions of this population towards their occupational safety. Drawing on in-depth interviews, this study examined the perspectives of healthcare professionals working in a large hospital during the pandemic, including their infection risks, the barriers or challenges to implementing their tasks, and the IPC measures to protect their safety and health. Healthcare professionals can offer insights into the practicalities of prevention and control measures, which have seen insufficient attention. This study aimed to inform the development of future recommendations to optimise compliance with appropriate use of these measures, and to improve the guidance to reduce their risk of the disease.

## Method

### Setting

The interviews were conducted from June to September 2020 at Ramathibodi Hospital, a public tertiary medical school hospital In Bangkok, Thailand. Since the beginning of the pandemic, the hospital provided COVID-19 testing and care for suspected and confirmed COVID-19 patients. The Virology unit at the hospital performed virology testing services for the hospital as well as other hospitals or clinics that sent their samples. An extended campus, Chakri Naruebodindra Medical Institute (CNMI), was subsequently designated to admit patients with confirmed COVID-19. In July 2021, about 300 medical workers at the hospital were infected during which the hospital provided care for approximately 1,000 COVID-19 patients, 350 patients in home isolation and 200 others waiting to be admitted for treatment (13).

### Ethical approval and consent to participate

Ethical approval was obtained from the Human Research Ethics Committee, Faculty of Medicine Ramathibodi Hospital, Mahidol University (approval reference: COA. MURA2020/828, 22 May 2020). All respondents provided written informed consent to be interviewed and interviews were audio-recorded at their places of work.

### Respondents and recruitment

Participants were recruited through a combination of purposive and convenience sampling. Initially, medical staff and health care workers who cared for suspected and/or confirmed COVID-19 patients in the hospital were invited to participate in an interview. Purposive sampling was used to ensure participants in a variety of roles in the hospital were interviewed. Additional participants were identified via a snowballing technique in various medical units, hence, we were able to reach those medical staff who may not have been captured in the initial phase. An attempt was made to recruit an equal proportion of hospital staff from different age groups so as to ensure a diverse set of experiences of working in the hospital setting.

### Data collection

Data collection was carried out using an iterative approach such that the research questions and interview question topics evolved as themes emerged from the data and as saturation was reached on a given subject area. Topics included: knowledge of, or experience with, COVID-19, risk factors and behaviours, perspectives on infection prevention and control strategy, and work activities and training (see Additional file 1). In-depth interviews began with a general discussion of their work activities during the COVID-19 pandemic.

The in-depth interview guide was developed based on the initial topics drawn from a recent qualitative evidence synthesis on barriers and facilitators of adherence to IPC guidelines for respiratory infectious diseases among healthcare workers (Table 1 (14)). Questions on the infection prevention and control strategy were designed based on WHO’s principles of COVID-19 IPC strategies in healthcare settings (Table 2 (9)). The guides were designed in English and translated to Thai. The guides, which included key topic areas and lists of suggested questions, were used in a flexible and iterative manner: interviewers would use the appropriate questionnaires to elicit information on the specific topic of interest. The guide was then piloted with the first recruited study participants to check for any miscommunication and revised as necessary. The number of interviews was determined by the point of saturation when no more novel information emerged.

**Table 1.** Facilitators and barriers to healthcare workers’ adherence with infection prevention and control (IPC) guidelines for respiratory infectious diseases. Adapted from Hougton et al. 2020 (14).

| Domains | Subthemes |  |  |
| --- | --- | --- | --- |
| 1. Organisational factors | Safety climate: what management support is needed to create a safe workplace (workload, staffing) | Communication of IPC guidelines: how best to communicate the guidelines | Availability of training programmes: what the training needs are and who should deliver the training |
| 2. Environmental factors | Physical environment: whether space, ventilation, and facilities are adequate | Availability of PPE: whether supply of PPE is adequate |  |
| 3. Individual factors | Individual knowledge: how knowledge of IPC guidelines and their importance influence adherence | Individual attitudes: how HCW perceive the value of IPC guidelines in protecting them, their families, and patients | Individual beliefs: how HCW's fears, concerns, and duty of care influence adherence to IPC guidelines |

**Table 2.** Principles of IPC strategies during health care when COVID-19 is suspected. Adapted from WHO Interim Guidance 29^th^ June 2020 (9).

| Strategies | Example measures |
| --- | --- |
| 1. Screening and triage for early recognition of patients with suspected COVID-19, and rapid implementation of source control measures | Establishing screening and triage, early identification and isolation of suspected cases, designated waiting area |
| 2. Applying standard precautions for all patients | Ensuring hand hygiene, respiratory hygiene; rational, correct and consistent use of PPE |
| 3. Implementing additional precautions | Managing isolation and cohorting of patients with suspected or confirmed COVID-19; contact and droplet precautions, airborne precautions |
| 4. Implementing administrative controls | Establishing sustainable IPC infrastructures and activities; ensuring adequate supplies of PPE; ensuring adherence to IPC policies and procedures |
| 5. Implementing environmental and engineering controls | Ensuring adequate ventilation and environmental cleaning; managing laundry, food service utensils and medical waste |

### Data processing and analysis

After respondents gave their consent, interviews were audio-recorded and subsequently transcribed verbatim and translated to English or summarised into detailed notes. The translated transcripts and notes were imported into NVivo version 12 (QSR International Australia) for qualitative content (thematic) analysis. An initial codebook was developed using established categories based on the original research questions and revised as new themes emerged from reoccurring data and during debriefs where the codebook was discussed and refined. The codebook was flexible, and the codes were reassessed during data collection and revised according to the emergence of novel themes. Transcripts were read several times and coded line-by-line using an inductive and deductive approach: the codebook used was initially based on the main research topics. Subsequently, during the process of coding, themes that emerged from the data were incorporated into the codebook.

### Findings

The findings presented are based on individual in-depth interviews with 23 healthcare workers in various roles including nurses, medical doctors, and laboratory technicians (Table 3). The main themes that emerged during the interviews were the participants’ perceptions of COVID-19 transmission risk at work, the various prevention measures and the challenges they faced while using these measures. There was also discussion about the need for training on the use of PPE and improving the IPC strategies in the hospital setting.

**Table 3.**
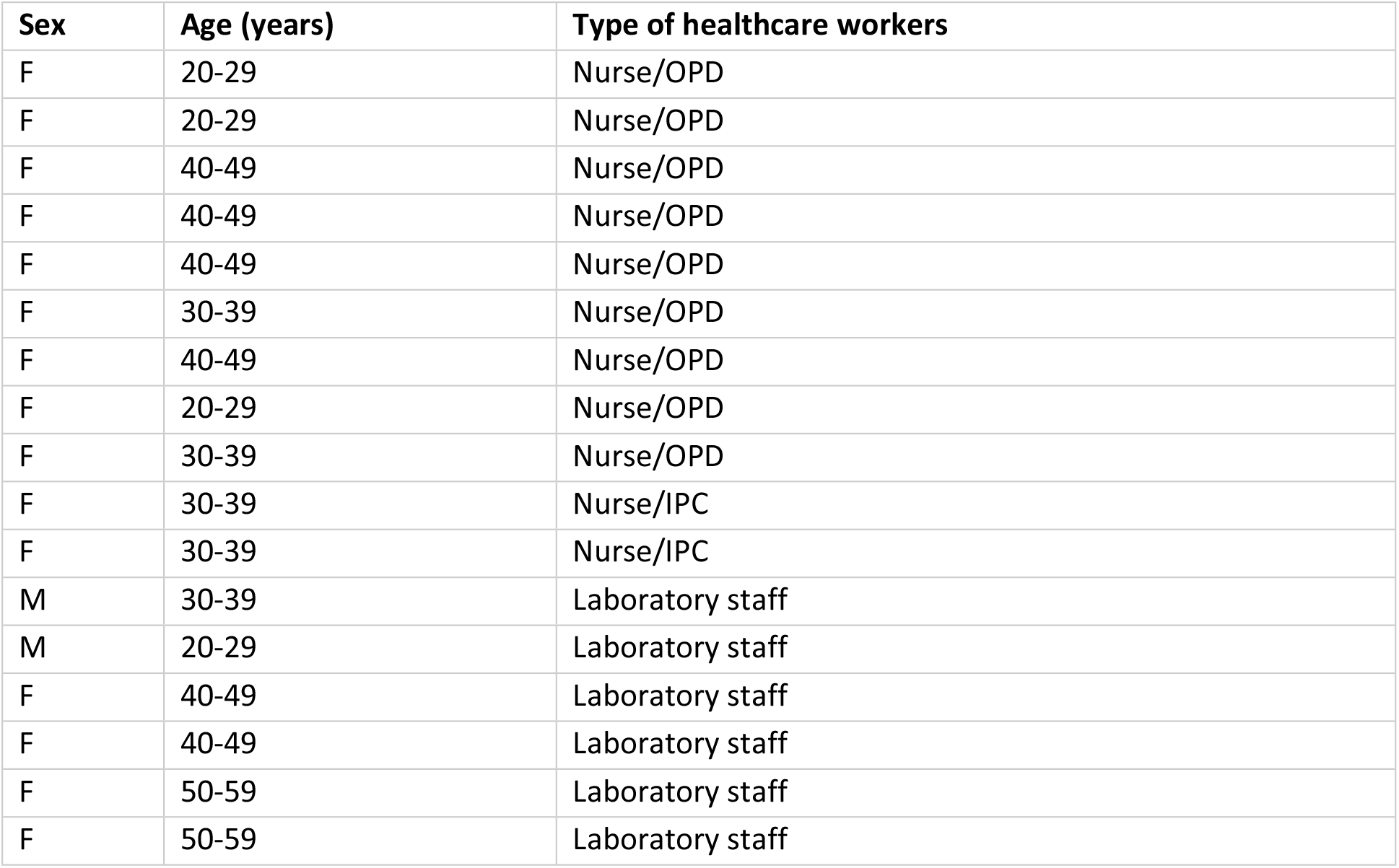
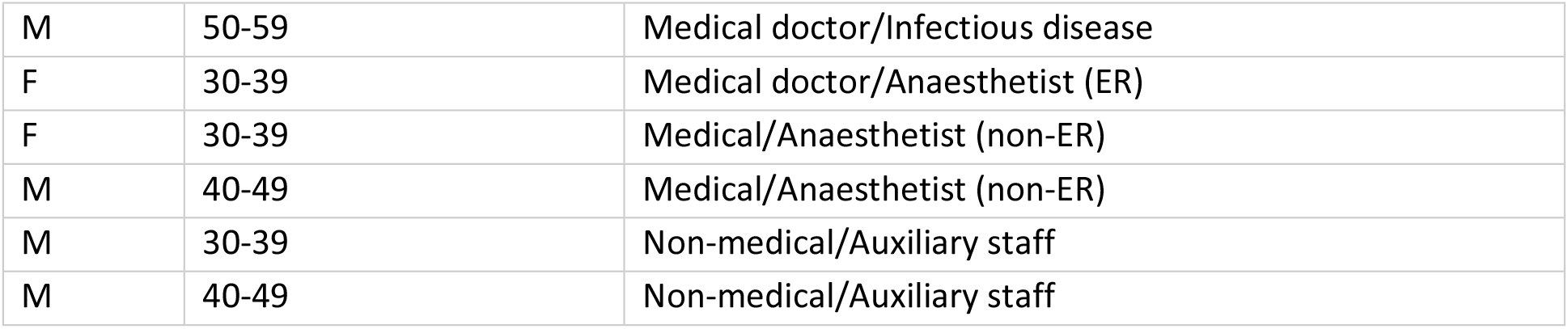
Characteristics of interview respondents. F=Female, M=Male, OPD=Outpatient Department, IPC=Infection Prevention and Control, ER=Emergency Room.

| Sex | Age (years) | Type of healthcare workers |
| --- | --- | --- |
| F | 20-29 | Nurse/OPD |
| F | 20-29 | Nurse/OPD |
| F | 40-49 | Nurse/OPD |
| F | 40-49 | Nurse/OPD |
| F | 40-49 | Nurse/OPD |
| F | 30-39 | Nurse/OPD |
| F | 40-49 | Nurse/OPD |
| F | 20-29 | Nurse/OPD |
| F | 30-39 | Nurse/OPD |
| F | 30-39 | Nurse/IPC |
| F | 30-39 | Nurse/IPC |
| M | 30-39 | Laboratory staff |
| M | 20-29 | Laboratory staff |
| F | 40-49 | Laboratory staff |
| F | 40-49 | Laboratory staff |
| F | 50-59 | Laboratory staff |
| F | 50-59 | Laboratory staff |
| M | 50-59 | Medical doctor/Infectious disease |
| F | 30-39 | Medical doctor/Anaesthetist (ER) |
| F | 30-39 | Medical/Anaesthetist (non-ER) |
| M | 40-49 | Medical/Anaesthetist (non-ER) |
| M | 30-39 | Non-medical/Auxiliary staff |
| M | 40-49 | Non-medical/Auxiliary staff |

### Perception of the risk of COVID-19 transmission

All respondents were aware of their risk of COVID-19 transmission at work. The risk was perceived to vary among different units or staff roles in the hospital. Respondents described that those assigned to treatment of COVID-19 patients were at increased risk. Nurses working at the outpatient department perceived that they might be at higher risk than the in-patient department (IPD) staff because they encountered more patients, either suspected or confirmed, and provided care for those patients regardless of their known status. Laboratory technicians who worked with COVID-19 samples reported high awareness of their risk but described that they are more prepared to protect themselves because of previous training and experience working with infectious disease samples.

HCW also identified places or areas that they perceived as higher risk for transmission in the hospital. Some respondents described a temporary OPD that was previously set up to screen and test suspected COVID-19 patients was somewhat unsuitable; they perceived an outdoor acute respiratory infection clinic (ARIC) to have lower transmission risk. One respondent compared risk of transmission between IPD and OPD whereby the latter was perceived to have higher risk of transmission for general staff and visitors. An IPC nurse described IPD to have higher risk for antimicrobial resistance as there were more patients with clinical symptoms who might receive treatment. Areas with high density and movement of people in the hospital such as waiting areas, screening points, and cafeteria, were also perceived to be of increased risk.

Respondents described concerns about the transmission risk related to certain medical procedures producing droplets and/or aerosols. This included performing intubation, suction, or resuscitation for patients in OPD and emergency units. One respondent compared risk of, and prevention used for, COVID-19 to tuberculosis in healthcare settings, for which the former is perceived to be likely transmitted by droplets and the latter by aerosols. Laboratory technicians also mentioned that some staff were concerned about probable contamination of COVID-19 sample delivery packages as they were delivered from multiple sites and might not be properly wrapped and boxed.

Concerns about potential for contracting COVID-19 infection at work were expressed by most respondents. Nine mentioned this concern related to subsequently infecting their family members and others in the hospital. One respondent described her concern about putting others she encountered, such as public transport users or neighbours, at risk. One respondent mentioned how her neighbour was aware of her work at the hospital and having less contact during the early pandemic. Some however argued that public spaces with high human density, such as public transport, markets or department stores, are of higher risk than the healthcare facility as the latter was better screened and managed.

HCWs also expressed concern over contingent cost from performing their work related to COVID-19. A few respondents described avoiding the use of public transport and needing to pay more for a private vehicle. One staff mentioned renting accommodation close by to avoid traveling and reduce the risk of infecting his family members at home. The majority of respondents described loss of their private or personal time, particularly with their families. One respondent described sending their children to stay temporarily with their parents. Some respondents described receiving compensation for their occupational risk from the government and hospital to compensate for their increased risk and additional working hours.

### Use of prevention measures

The COVID-19 prevention measures that respondents reported using were N95 and surgical face masks, face shield, hand washing, PPE, screening for risk factors and symptoms (by taking temperature and/or contact history), social or physical distancing, ventilation, use of negative pressure rooms, and self-quarantine or isolation.

Respondents discussed how wearing face masks and washing their hands were the most important measures when working in the hospital. PPE was identified as crucial for staff (particularly doctors and nurses) when testing suspected cases or caring for confirmed patients, and handling samples (for laboratory staff). Use of respirators was reported to be essential for anaesthetists and staff in the emergency room (ER) to perform medical procedures. All categories of staff reported these personal protection measures made them feel safer at work.

Cleaning and sanitising the environment was described to be a necessary measure in the hospital, particularly for units which have suspected and confirmed COVID-19 patients. Staff mentioned a regular schedule for cleaning medical and auxiliary areas and changing patient’s bed sheets. A group of nurses in the ARIC reported ventilation to be among the most important measures, especially during periods when a high volume of patients visited the clinic for COVID-19 testing.

Nurses who were tasked to care for patients with confirmed COVID-19 and/or worked in a COVID-19 ward mentioned about having to self-quarantine for 14 days after their shift to self-monitor and reduce the risk of transmission to their co-workers when returning to their ward.

### Challenges using COVID-19 prevention measures

Respondents discussed several reasons for their use and non-use of COVID-19 prevention measures. Most respondents described having high awareness about using the prevention measures and having received appropriate guidance on how to apply them according to their roles. Challenges of using certain measures were described; for example nurses described taking off the PPE as being a difficult process and they needed help from a colleague to properly remove the equipment and avoid self-contamination. One respondent reported that the appropriate use of PPE may depend on experience of the staff and the material of the PPE; material with a harder texture was perceived as being more difficult to remove properly. For masks, respondents described the importance of doing a fit test once or twice a year to ensure correct use; some mentioned also using plastic tape to ensure full protection from a mask. A few respondents reported that wearing an N95 mask and face shield reduced their ability to see when performing certain medical procedures such as injection or drawing blood. A few respondents mentioned that they were unsure whether UV light can disinfect face masks and for how long.

> “*It’s really important to always do a fit test … not only one time a year but regularly, especially when dealing with airborne transmission. Tuberculosis, for example. Fit test for N95, which brand or type, you need to check and be aware of this to be safe*.” IDI with IPC nurse

The second challenge was the physical setup. Respondents described inadequate space for staff to socially distance themselves at work, particularly where more staff worked full-time in a small area. Some also mentioned that patients were sometimes unable to maintain two-metre physical distancing in a small or crowded space such as a waiting area or restroom. In addition, respondents discussed a lack of necessary material and equipment such as N95 masks and PPE, especially during the first few weeks of the pandemic. During shortages of protective equipment, respondents described that high-risk HCW were prioritised, specifically those who had direct contact with confirmed patients.

> “*We were fortunate that the number of patients did not outnumber our capacity, our equipment. So as long as the equipment is sufficient, we are OK. Patients were very well-taken care off. I could not imagine what would happen if the patients outnumbered the negative pressure rooms we have. We might be in trouble … even though we are prepared, there could be situations that might put us at risk*” IDI with medical doctor

Some respondents also described their use and non-use of certain measures such as masks related to the hospital infrastructure. For example, staff stationed at an ad-hoc outdoor ARIC found it uncomfortable to wear a face mask and a face shield for long hours in hot and humid weather. A few nurses also mentioned that it was difficult to communicate with patients through the mask, face shield, and an acrylic barrier. Use of telecommunication was described by most staff to be effective for interacting with patients while keeping distance; some however mentioned that they communicated less effectively with patients given the physical barriers. One respondent also reported observing a patient feeling uncomfortable in an outdoor waiting area for an extended period of time. Nevertheless, better ventilation at the outdoor ARIC was perceived to be positive by an IPC nurse who expressed concern about overcrowded, indoor environments in the hospital.

Respondents discussed the importance of cooperation from others including co-workers, patients, and visitors to effectively implement the measures. The first challenge was related to patients’ behaviour. Staff reported that some patients who visit their unit failed to report their risk of COVID-19 infection, either because they were unaware of their own risk or they felt stigmatized to disclose this information to the staff during screening (or both). Some described that this caused delay in giving the right treatment to the patient as well as increasing the transmission risk for the patient and the staff themselves. For example, a patient reported having had contact with a high-risk person days after she first visited the unit for her respiratory symptoms, and during which staff implemented a minimal level of prevention. Staff discussed how the patient might have received more appropriate care and experienced a quicker recovery if she had reported her travel and contact history when initially assessed. Staff who provided care for the patient reported feeling anxious and needing to isolate herself from her colleagues and family members for fear of inadvertently transmitting the disease to them. For another patient, staff described how they only agreed to disclose his information to the doctor in a closed room to maintain his privacy.

Staff described various ways to deal with the challenges. Respondents mentioned prolonging use of N95 and surgical masks, purchasing masks for their own use, wearing two masks in a double layer, and use of UV light to disinfect masks. Hospital telephones or chat applications were used to reduce the time physically interacting with patients and colleagues and reducing the risk from contaminated documentation. An acrylic box which was used to cover patient’s head and neck was mentioned to be effective and resource-efficient when performing intubation, resulting in lower use of masks and face shields during a shortage. Instructions on how to use PPE were provided to staff so that they were able to perform their tasks; nurses mentioned that a newly established swab unit in the ARIC had helped reduce their need to use PPE when testing for COVID-19.

Donations was described as an alternative to supply necessary material and equipment, and for other additional support such as meals. Respondents reported that these had helped to alleviate the period of supply shortage, although important equipment such as masks and PPE needed to be assessed for quality before use. Other measures, such as rescheduling appointments with non-emergency patients were implemented to reduce the number of visitors in the facility. OPD nurses also described specifically designating the task of caring for patients with suspected COVID-19 to one staff per shift or unit to minimize staff’s contact with the patients and therefore reduce the risks for other patients receiving care in the unit.

### Implementation of infection prevention and control measures

Several factors were identified for effective implementation of COVID-19 IPC measures: training and provision of information, availability of prevention equipment, clear guidelines and workflow, collaboration among units, and effective communication with patients.

#### Training and provision of information

The majority of the interviewed staff felt they were well informed about the information regarding COVID-19 disease and had received necessary training for infection prevention and control. Interviews with IPC nurses identified that prioritised training included fit testing for masks and powered air purifying respirators (PAPR), for which only the emergency unit was usually prepared; training was provided to other units after the initial outbreak. Training on the use of PPE was also discussed; respondents described that the terms used to identify which types of PPE were needed had been adapted for COVID-19 related tasks. Formal PPE training for ARIC staff was prioritized. Lab staff described that they had received multiple training sessions as part of their job, and hence felt that they were well equipped to safely carry out COVID-19 related tasks.

Managing the concerns of HCW was perceived to be a challenge, particularly at the beginning for the IPC team. They described COVID-19 as an emerging disease and compared it to the early years of the AIDS pandemic when the disease was perceived to be unknown and stigmatizing. A few staff who had had prior experience with MERS described that they felt prepared for this pandemic; however, some respondents felt that the training on PPE could be provided for a wider group of staff because presumptive COVID-19 patients may be encountered by general OPD nurses and staff. Training targeting non-medical staff was also suggested to be important because they also had risk of exposure in the hospital setting. These included cleaners, receptionists, and security guards who may also not be Thai-nationals.

For this training, nurse respondents described the benefit of different learning tools, such as educational videos and posters, to help remind the staff when performing their tasks. This was described as helping remind staff about the steps for how to properly don or doff PPE; thus reducing their risk and anxiety about self-contamination.

#### Use and availability of prevention equipment

When asked about the long-term plan for IPC, respondents mentioned that preparing the staff and keeping the prevention equipment available is important to prepare and alleviate staff’s concern about their risk. IPC nurses also described demonstrating use of appropriate prevention measures and helping to care for patients with confirmed COVID-19, made staff feel more confident and prepared to take on the job, and was described as a way to give them moral support. In addition to the use of prevention measures, the IPC nurses also reported that HCW need to monitor themselves under the current working circumstances and should report for testing if they had symptoms, to avoid risk of transmission and reduce concerns among staff. One respondent also suggested increasing awareness about some prevention measures, such as correct elevator use, social distancing and hand hygiene, in the hospital.

#### Collaboration among units

Collaboration among units in the hospital was reported to be an important factor to carry out this training. Respondents described how various HWC contributed to each part of COVID-19 training: : medical doctors provided knowledge about the disease and how it is transmitted and demonstrated how to do a swab test; IPC nurses provided guidance on how to use the prevention equipment. IPC nurses also described working with the virology laboratory to determine how swab samples should be collected and packaged before sending them for analysis.

> “*In terms of communication, we really needed collaboration from many departments. Medical doctors could explain to staff about the disease and its transmission route … IPC nurses would give training on how to use prevention measures. Fellows would need to demonstrate how to do a swab test. We also needed to communicate with laboratory staff about how the samples will be taken and they informed us about how to deliver them … training was clear and well-delivered so that most nurses are able to do a swab test now*.” IDI with IPC nurse

Communication channels among HCW were set up to keep staff informed; however, respondents sometimes felt exhausted from changes in updated guidance and recommended practice, particularly early in the pandemic. Respondents described being aware of the changing nature of knowledge and practice for a newly emerging disease. Some senior staff emphasized giving clear instruction and updated information as an important way to avoid fatigue and confusion at work. IPC nurses also described simplifying and shortening some guidelines to make them more accessible and easier to understand.

#### Clear guidelines and workflow

Interviews with HCW found that updating protocols contingent on new information about the disease was a challenge; for example, what symptoms should trigger screening or which countries are high risk. Respondents described the importance of precision when screening eligibility of suspected patients for government-subsidized testing. This had caused difficulty for OPD staff who might not be aware of these constantly-updating criteria, such as addition of high-risk symptoms or countries. A nurse respondent who was tasked with managing the updated processes for screening and case investigation reported feeling pressured to give constant guidance to other hospital staff and monitoring their adherence to the guideline.

Respondents found that performing a demonstration of the written guidelines enabled staff to get comfortable with the protocols and steps such as screening and triage, and using PPE. Interviews with IPC nurses described that this type of demonstration enabled staff to practice in, and be prepared for, real-life situations. One respondent also mentioned that this allowed staff to work together to adapt the guidelines into practice based on the context of each ward or unit, for example, determining specific tasks and the number of staff needed based on the resources and capacity.

> “*At the infection control unit we normally provide training or suggestions to staff and not always by the book. We also observed their working environment and asked about their needs … to make them feel less worried, make ourselves part of their work, that we were in this together and also let the staff participate in adapting the guideline for their own use*” IDI with IPC nurse

#### Communicating with patients

Staff described communicating with patients to be a challenge; they needed to provide accurate information to patients with a positive COVID-19 test in a clear and empathetic way. Making notes of those conversation with patients was described to be useful to keep track of the patient’s concerns and emotional state. This was also mentioned as a way to provide patients with better care and to substitute for limited contact staff may have with patients. Respondents also reported doing so in different languages, particularly in Chinese and English, in the early phase of the pandemic before state quarantine was implemented. An IPC nurse described that they were aware of the importance of these tasks; however, they felt overwhelmed by the number of phone calls and were unable to continue their training work. Setting up a call centre with staff tasked to professionally communicate and allocate time for the task was mentioned to be a solution preferred by the staff.

## Discussion

This study used in-depth interviews to explore the perceptions of healthcare workers about perceived risk, use of prevention measures, and implementation of IPC measures to prevent COVID-19 transmission in a hospital setting in Thailand. Our findings identified the risks and concerns of healthcare workers at their work, the need for adequate training and appropriate protection equipment, and the facilitators and barriers to IPC measures in the hospital setting.

### Perceived risk, awareness, and concerns of healthcare workers

Our findings show that HCWs were aware of and used prevention practices in healthcare facilities; however, these were not always easy to implement in daily work routines. Respondents perceived their risk in healthcare settings with regards to their roles. Anaesthetists and ER staff were perceived to be at an increased risk compared to other medical staff because of the medical procedures they performed on both confirmed and unconfirmed patients. Similarly, it has been shown that healthcare workers involved in procedures which generate aerosols had the highest occupational risk from COVID-19 (15). Some respondents also described the risk based on location in the hospital and how it may affect their level of protection used; risk of infection was perceived to be higher in an outpatient unit because the unit provided care for a mixed group of patients whose COVID-19 status was unknown. In addition, their use of masks, physical barriers made of plastic or acrylic, and telecommunication have reduced their ability to provide empathetic care to the patients. This has been seen elsewhere with, for example, one nurse assistant infected from performing routine medical care on a dengue patient who was later diagnosed with COVID-19 (16). For tuberculosis, the use of masks and respirators had alienating or depersonalizing effects on medical staff and their patients (17).

Similar to our findings, several concerns about infection and transmission risk of COVID-19 among healthcare workers were identified elsewhere. Medical staff were concerned about limited access to appropriate PPE, testing, up-to-date information and communication, ability to provide competent medical care if HCWs were deployed to a new area (such as non-ICU nurses having to function as ICU nurses), and increased support for other personal needs (such as meals, accommodation and transportation) (18). Being exposed to COVID-19 at work and transmitting it to those at home were the main causes of anxiety among HCWs (19), especially when the workers live within a multigenerational household with elderly relatives (20). HCWs may also be negatively affected by economic and social impacts from COVID-19 public health measures (21): in Vietnam, higher cost of living and decreasing income were the main causes of concern for frontline HCWs (22). This implies that HCWs and other staff in healthcare facilities should be prioritised for prevention interventions, including COVID-19 vaccination, to reduce their risk of infection and prevent transmission in healthcare settings (2). A provision of support package on well-being (23) and mental health support to HCWs (24, 25) were recommended to help HCW cope with increased stress, anxiety, depressive symptoms and enhance the capacity of HCWs during the pandemic. This is particularly necessary for those in low-resource settings in this region (22), including community-based facility workers in Cambodia (26), Thailand (27), Vietnam (28).

### Physical environment and availability of protection equipment (environmental factors)

As identified by respondents, insufficient physical distancing between patients and co-workers, and indoor settings, were identified as among the most important risk factors by healthcare workers (29). Respondents also experienced discomfort from the use of N95 masks, face shields, and coveralls, especially when used outdoors in hot and humid weather. In the UK, use of PPE has been shown to cause heat stress and may negatively affect performance, safety and well-being of HCWs during pandemics (30). Adaptation of physical space and working environment is therefore crucial in response to some implementation measures, including weather conditions and social distancing (31). A recent study on an outbreak investigation reported transmission among HCWs at a quarantine facility in Thailand, and suggested use of thorough prevention measures and setting up private living quarters to reduce exposure risk for HCWs whilst performing their clinical work (32). In Malaysia, HCWs were also found to be particularly at risk at work when COVID-19 was not suspected in patients and insufficient PPE was worn (33), recommending that occupational health check-ups should be conducted to obtain information about the epidemiology of COVID-19 among HCW. Employment of mobile phone application technology to assist patients to self-identify their symptoms was also found to be effective in a Thai hospital setting (34).

Insufficient supply of prevention materials is a barrier to adequate or appropriate use of COVID-19 prevention measures, such as N95 masks and PPE. From the interviews, the acute shortages in medical supplies like PPE and equipment such as ventilators, were reported to be one of the main challenges early during the pandemic. Respondents reported using coping strategies in using them, however, these strategies may reduce the preventive effectiveness or increase chances for self-contamination. IPC nurse respondents highlighted the significance of improving the management of the supply to ensure that all medical professionals have enough resources to do their jobs as well as protect themselves from the disease. In many countries, hospitals faced limited operational capacity during the beginning of the pandemic due to a large demand shock triggered by acute need for healthcare supplies and health equipment, particularly in low-income countries (35). A recent survey among HCW suggested that adequate supply of PPE and emergency preparedness policies need to be put in place to alleviate concerns and anxiety of HCWs during the pandemic without compromising the safety of workers and patients (36).

### Training, communication, and workplace management (organizational factors)

The findings also show that training is crucial to ensure that staff are capable and familiar with practicing IPC measures. Some respondents highlighted that training should also include auxiliary staff who are not healthcare workers, such as cleaners, because they often assist in cleaning contaminated areas in the facility. More importantly, IPC nurses emphasized the importance of motivating HCWs to implement the prevention measures; support from and collective adherence to such practices among trainers and other colleagues are crucial to positively influence HCWs’ motivation. A recent review on adherence to IPC guidelines for respiratory infectious diseases among HCW also highlighted the need to include all staff including cleaning and kitchen staff in health facilities when implementing such measures (14). In Thailand, a recent national survey reporting overuse and underuse of PPE among HCWs during the pandemic in Thailand highlighted that training programmes should be provided and continue to ensure appropriate PPE practice in healthcare settings (37). In addition, training HCWs on contact tracing and testing among high-risk individuals, supported by the use of a mobile application for reporting, was found to help mitigate COVID-19 transmission in Cambodia (38). Knowledge about the severity of the disease increased Vietnamese frontline workers willingness to be vaccinated against COVID-19 (39). Adequate training and appropriate provision of PPE are therefore critical to encourage implementation of prevention measures and reduce differential access to adequate PPE.

From the interviews, HCWs valued clear and comprehensive guidelines to be able to do their work effectively; however, some respondents reported feeling overwhelmed by multiple tasks and practices during the early spread. IPC nurse respondents also highlighted that ambiguous and repeatedly changing guidelines may lead to additional workload and work fatigue among HCWs because they needed to take PPE on and off, do additional cleaning, or rearrange their staff and physical space in their unit. Studies on the 2015 MERS outbreak in South Korea also showed that ambiguous and frequently changing guidelines (40), and nonstandardised protocols (41) resulted in HCW’s confusion and burnout. Practical steps taken by IPC nurses suggested that involving staff in adapting the guidelines to the context of each unit, and communicating them in an accessible and transparent manner are effective measures to encourage HCW’s adherence to the guidelines.

## Strengths and limitations

To our knowledge, this is the first study that has used qualitative research methods to specifically address perceptions of healthcare workers during the COVID-19 pandemic in a hospital setting. The findings are mainly drawn from self-reported information and might be subject to desirability bias, however, observations and a previous survey (12) provided additional information in the context of healthcare setting and on sensitive topics, such as anxiety. Most respondents were female, which reflects the proportion of female nurses in the health facility, and were a diverse group of medical staff, drawn from a range of units and roles. Two non-medical staff were interviewed; however, findings regarding their perception may be less diverse because the study did not include other groups of staff such as cleaning and security personnel. High awareness and preparedness of respondents in the study may also be related to the advanced health facility they were working in, which may limit the generalisability of the findings to other facilities. Further research should ideally explore the experience of HCWs in a variety of health facilities, particularly in resource limited settings.

## Conclusion

Healthcare workers perceived themselves as at increased risk of COVID-19 infection in hospital settings during the pandemic. Several factors influenced the use of multiple prevention measures: concerns about infection, availability of consumables and equipment, barriers to work performance, and physical limitations in the hospital setting. Adequate training, clear guidelines, streamlined communications, and management support are crucial to encourage appropriate use of, and adherence to, implementation of IPC measures among HCW. Factors to effectively implement prevention measures and practical guidelines may vary between health facilities. Further studies should thus explore the perceptions and experiences of HCW about how best to protect HCW and patients from COVID-19 transmission in a variety of types of health facility during the COVID-19 pandemic.

## Data Availability

The data on which this article is based cannot be shared publicly due to confidentiality of the individuals who participated in the study. The data are available upon reasonable request to the Mahidol Oxford Tropical Medicine Research Unit Data Access Committee complying with the data access policy (https://www.tropmedres.ac/units/moru-bangkok/bioethics-engagement/data-sharing/moru-tropical-network-policy-on-sharing-data-and-other-outputs) for researchers who meet the criteria for access to confidential data.

## Abbreviation

ER: Emergency Room
HCW: Healthcare worker
IPC: Infection prevention and control
KAP: Knowledge, attitude, and practice
OPD: Outpatient Department
PPE: Personal protective equipment
UV: Ultraviolet
WHO: World Health Organization

## Acknowledgements

The authors would like to thank the respondents who participated in the study and took time to share their experiences and opinions with members of the research team. We would also like to thank a team of staff from Ramathibodhi hospital, including Keetakarn Taleangkaphan, Saowalak Khongsamrit, Sumawadee Sakuntaniyom, and Suthunyapat Maleehual who supported the ethics application and data collection. We wish to acknowledge Worarat Khuenpetch and Arissara Krataijun for their help with data management.

## Authors’ contributions

RJM RRM MJ designed and conceptualised the study

MJ collected the data.

MJ PC analysed the interviews and wrote the first draft of the manuscript.

RJM RRM supervised the study.

All authors contributed to and approved the final version.

## Competing interests

None

## Funding

This research was funded in whole, or in part, by the Wellcome Trust [Grant number 220211]. For the purpose of open access, the author has applied a CC BY public copyright licence to any Author Accepted Manuscript version arising from this submission.

## Additional file

In-depth interview guide

## Notes

### Competing Interest Statement

The authors have declared no competing interest.

